# Examining the Unit Costs of COVID-19 Vaccine Delivery in Kenya

**DOI:** 10.1101/2021.11.01.21265742

**Authors:** Stacey Orangi, Angela Kairu, Anthony Ngatia, John Ojal, Edwine Barasa

## Abstract

**Introduction:** Vaccines are considered the path out of the COVID-19 pandemic. The government of Kenya is implementing a phased strategy to vaccinate the Kenyan population, initially targeting populations at high risk of severe disease and infection. We estimated the financial and economic unit costs of procuring and delivering the COVID-19 vaccine in Kenya across various vaccination strategies.

**Methods:** We used an activity-based costing approach to estimate the incremental costs of COVID-19 vaccine delivery, from a health systems perspective. Document reviews and key informant interviews (n=12) were done to inform the activities, assumptions and the resources required. Unit prices were derived from document reviews or from market prices. Both financial and economic vaccine procurement costs per person vaccinated with 2-doses, and the vaccine delivery costs per person vaccinated with 2-doses were estimated and reported in 2021USD.

**Results:** The financial costs of vaccine procurement per person vaccinated with 2-doses ranged from $2.89-$13.09 in the 30% and 100% coverage levels respectively, however, the economic cost was $17.34 across all strategies. Financial vaccine delivery costs per person vaccinated with 2-doses, ranged from $4.28-$3.29 in the 30% and 100% coverage strategies: While the economic delivery costs were two to three times higher than the financial costs. The total procurement and delivery costs per person vaccinated with 2-doses ranged from $7.34-$16.47 for the financial costs and $29.7-$24.68 for the economic costs for the 30% and 100% coverage respectively. With the exception of procurement costs, the main cost driver of financial and economic delivery costs was supply chain costs (47-59%) and advocacy, communication and social mobilization (29-35%) respectively.

**Conclusion:** This analysis presents cost estimates that can be used to inform local policy and may further inform parameters used in cost-effectiveness models. The results could potentially be adapted and adjusted to country-specific assumptions to enhance applicability in similar low-and middle-income settings.

## INTRODUCTION

COVID-19 cases and deaths have been on the rise globally and locally since the initial case was reported in Kenya on 13^th^ March 2020. As of October 28 2021, there have been 252,938 confirmed cases and 5,266 reported deaths in Kenya [1]. COVID-19 vaccines are considered a path out of the pandemic. Research and development of COVID-19 vaccines has been prioritized globally and accelerated, with 194 vaccines in pre-clinical development and 128 vaccines in clinical development as of late October 2021 [2]. At the time of writing this, 7 vaccines have currently been deemed safe, of good quality and effective, and have been approved for use by the World Health Organization [3].

Kenya’s COVID-19 vaccine deployment plan targets to vaccinate 50%-70% of its adult population by end of June 2022 while prioritizing those who are at an increased risk of severe disease and infection [4]. The country’s primary procurement source is the COVID-19 Vaccines Global Access (COVAX) Facility. However, given vaccine availability constraints with the COVAX facility, the country is pursing alternative vaccine supply and procurement options that include donations from other countries, the African Unions (AU) African Vaccine Acquisition Task Team (AVATT) mechanism, as well as direct procurement from vaccine manufactures. As of October 28 2021, a total of 5,153,667 vaccine doses had been administered [5]. Only 13% of the adult population has received their first dose while 5.7% have received their second dose and are fully vaccinated [5]. The main vaccine that has been deployed is the Oxford/AstraZeneca (2-dose per person), however, other vaccine formulations have been procured and are soon to be deployed.

Evidence on the costs of delivering COVID-19 vaccines are useful for several reasons. First, the financial costs of delivery are useful inputs in decisions and planning about resource mobilization for the vaccine programme. Second the financial costs of delivery provide inputs for assessments of budget impact and affordability of vaccination deployment scenarios, and hence can inform vaccine priority setting decisions. Third, economic costs of vaccine delivery are useful for parametrizing vaccine cost-effectiveness models that will provide evidence on the value for money of different vaccination strategies, and threshold prices that can guide price negotiations to improve the efficiency of procurement. This study aims to estimate the financial and economic unit costs of procuring and delivering the COVID-19 vaccine in Kenya across various vaccination strategies.

## METHODS

### Study setting

Kenya is a low-and middle-income country with a GDP per capita of $1,838.21 [6]. The total population in 2019 was estimated at 47.5 million with a predominant youth population; 48% of the population are aged 18years and below [7]. The country has a devolved system of governance characterized by a national government and 47 semi-autonomous counties [8]. Healthcare service provision is provided through a pluralistic system in both the public and private sector. The public sector is organized into four main tiers 1) community health services 2) primary care 3) county referral services 4) national referral services [9].

### COVID-19 vaccination plan in Kenya

The government of Kenya has a phased approach of vaccine deployment that prioritises the population receiving the vaccine based on their vulnerability, vaccine availability and the health system capacity [4]. Phase 1, between March-June 2021, was the initial roll-out planned during limited vaccine supply; This phase targeted front-line workers and older persons (aged 58years and above) [4]. Subsequently, phase 2 was planned for July-December 2021 when a larger number of vaccine doses would be available [4]. In addition to the priority group not reached in phase 1, phase 2 would target 1) those aged 50 and above 2) Individuals 18years and above with co-morbidities and those living with disabilities 3) Individuals working in the hospitality and transport sectors 4) Individuals in congregate settings [4]. Lastly, phase 3 planned for January to December 2022 would focus on open mass vaccination [4]. Vaccination was planned to be administered in identified COVID-19 vaccine provider sites from public and private sectors, as well as progressively including targeted outreaches, and eventually campaigns [4]. The COVID-19 vaccine is distributed from the national stores to the regional depots, and finally to the vaccination health facilities, and follows existing distribution patterns.

### Costing approach

This study employed a health-system’s perspective. An activity-based costing approach was used to determine the incremental costs of the introduction of COVID-19 vaccines in Kenya, in line with guidelines for estimating the costs of new vaccines introduction [10,11]. The time horizon was one year.

### Costing scenarios and assumptions

Four different vaccination coverage levels were considered; 30%, 50%, 70%, and 100% of adult Kenyan population receiving two-doses of COVID-19 vaccine at the health facilities, within a one-year period. The selected vaccination scenarios reflect the evolution of Kenya’s COVID-19 vaccine deployment strategy. The lowest coverage scenario (30%) represents the initial coverage target of Kenya’s COVID-19 deployment plan at a time when the country was less optimistic about the availability of vaccines. Given the unpredictability of vaccine supply, we have assumed that this low coverage scenario might still be reflect what may happen in practice, despite subsequent revisions of the vaccination strategy to revise the coverage target upwards. The 50%, 70% and 100% coverage scenarios reflect targets in subsequent iterations of the deployment plan.

The cost analysis made some assumptions shown in Table 1. First, a 2021 target population was used which was derived by adjusting the 2019 population reported in the census with a 2.2.% population growth rate [7,12]. In terms of vaccine and related supplies, a wastage rate of 10% was used for both syringes and vaccines respectively [13]. An assumption was made that vaccines procured by the COVAX facility and through bilateral negotiations would be classified as economic costs and cover 20% and 5% of the population respectively, regardless of the vaccine strategy. This was assumed based on COVAX strategy to cover 20% of the population and the existing ratio of procured vaccines in the country from COVAX and bilateral negotiations. Lastly, the primary costing analysis assumed 2 doses per person.

**Table 1:** Costing assumptions.

|  | Estimate | Data Source |
| --- | --- | --- |
| **Demographics** |  |  |
| Target adult population (2021) | 26,781,011 | 2019 census report[7] |
| Population growth rate | 2.2% | 2019 census report [12] |
| **Vaccine strategies** |  |  |
| 30% coverage | 17% of vaccines procured by government<br>83% of vaccines procured by COVAX and through bilateral negotiations | Assumption: Vaccines procured by COVAX and through bilateral negotiations cover 20% and 5% of the population, regardless of vaccination strategy |
| 50% coverage | 50% of vaccines procured by government<br>50% of vaccines procured by COVAX and through bilateral negotiations |  |
| 70% coverage | 64% of vaccines procured by government<br>36% of vaccines procured by COVAX and through bilateral negotiations |  |
| 100% coverage | 75% of vaccines procured by government<br>25% of vaccines procured by COVAX and through bilateral negotiations |  |
| Vaccine and related supplies characteristics |  |  |
| Vaccine wastage rate | 10% | [13] |
| Syringes wastage rates | 10% | Interview with national level official |
| Number of doses on schedule | 2 doses per person |  |
| Add-on vaccine procurement costs |  |  |
| Freight and insurance fees | 3.0% | EPI |
| Handling fees | 4.0% |  |
| Clearance charges | 0.1% |  |
| Import declaration fees | 2.0% |  |
| Railway development levy | 3.5% |  |

### Unit costs

We estimated the following unit costs (Figure 1):

- Cost of vaccine procurement per person vaccinated with two-doses
- Cost of vaccine-related supplies procurement per person vaccinated with two-doses
- Cost of vaccine delivery per person vaccinated with two-doses

**Figure 1:**
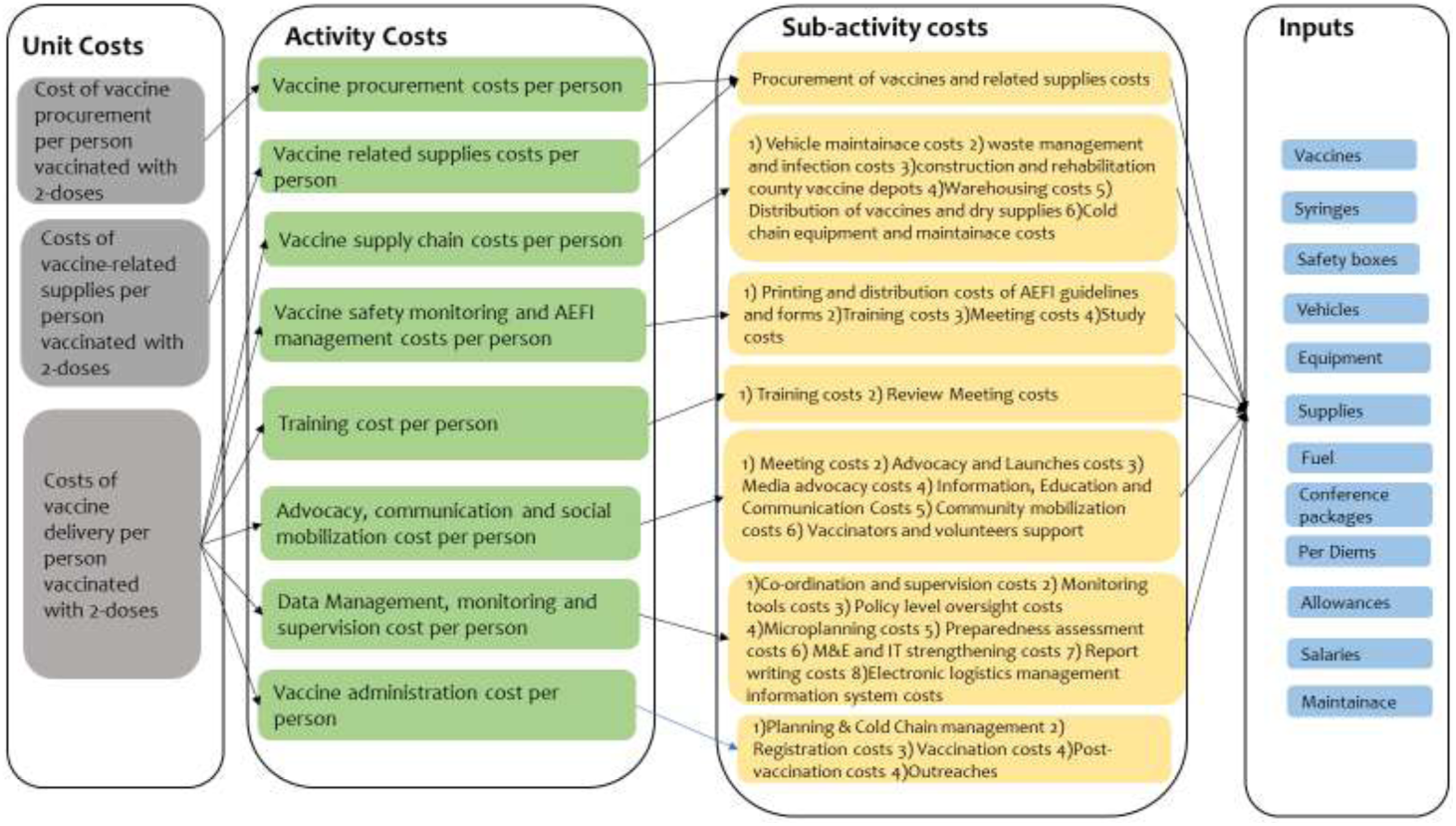
Identification of COVID-19 costs.

Procurement costs refer to the purchasing costs of the COVID-19 vaccine and its related supplies, accounting for the importation and clearance costs into the country. The vaccine procurement cost per person vaccinated with two-doses and the cost of vaccine related supplies procurement per person vaccinated with two-doses was estimated by dividing the total costs of vaccine procurement and the total costs of vaccine-related supplies procurement respectively by the total number of people targeted for vaccination.

Delivery costs refer to the costs of activities required to supply and distribute the vaccine from the national level to the health facilities, as well as activities that ensure the safe administration and uptake of the vaccine. Cost of vaccine delivery per person vaccinated with two-doses was estimated by obtaining the total costs of 1) vaccine supply chain 2) vaccine safety monitoring and adverse events following immunization (AEFI) management 3) training 4) advocacy, communication and social mobilization 5) data management, monitoring and supervision, and 6) vaccine administration and dividing this by the total number of people targeted for vaccination.

### Cost components

The different activities identified in the delivery and implementation of the COVID-19 vaccine were categorized under 7 key components; 1) Vaccine and related supplies procurement costs 2) Vaccine supply chain costs 3) Vaccine safety monitoring and AEFI management 4) Training 5) Advocacy, communication, and social mobilization 6) Data management, monitoring and supervision 7)Vaccine administration during service delivery. Under each cost component, there were identified sub-activities and inputs that were costed. This is illustrated in Figure 1. The cost components are further detailed below.

#### i) Vaccine and related supplies procurement

This included the costs to procure the vaccine and its related supplies. Vaccine procurement base costs was estimated at US$7 per dose, which is the country’s procurement cost from the COVAX facility. Additionally, freight and insurance fees, handling costs, clearance charges, import declaration fees, and railway development levies were added as a percentage of the base cost of vaccine and related supplies costs. The number of doses to be procured was estimated based on the number of doses to be given and adjusted for the wastage rates.

#### ii) Vaccine supply chain costs

Vaccine supply chain costs included the costs of distribution of the vaccines and related supplies from the national depots, through the regional depots and eventually to the healthcare facilities. This included capital costs such as the procurement of cold chain equipment, waste management and control equipment, vehicles for last mile distribution, and renovations of county vaccine stores. Recurrent costs included vehicle maintainace costs, waste management and infection control supplies, warehouse costs, transport costs of vaccines and dry supplies, and cold chain preventive maintainace visits.

The number of resources, frequency of distribution were estimated at a national level and the total amount of resources required estimated using ingredients costing of the different inputs such as supplies, utilities of the warehouses, fuel, per diems of the drivers and other staff involved in the distribution process, and staff salary costs. The capital investments were valued as a shared input with other vaccines in the Expanded Programme on Immunization (EPI). Therefore, in order to account only for the incremental costs, apportioning of the capital costs was done by allocating a fraction of distribution costs to COVID-19 vaccine based on all vaccine doses given in Kenya in one year.

#### iii) Vaccine safety monitoring & AEFI management

Vaccine safety monitoring and AEFI management included sub-activities to support AEFI field investigation, sensitization meetings on vaccine safety and risk management, pharmacovigilance sensitizations and supervision support, quality assurance for supply and vaccine safety, printing of national AEFI guidelines and forms and vaccine safety study. Costs of these sub-activities included conference packages, printing, fuel, airtime, per diem allowances for travel and lunch, and salaries.

#### iv) Training

This included the initial training of logisticians at a national and county level, initial training of sub-county health management team (SCHMT) supervisors, and the initial training of vaccinators and volunteers. Additionally, review meetings of the vaccination program at a national, county and sub-county level were included. For each of the trainings and review meetings the estimated costs included conference packages, per diems for participants and facilitators, lunch costs, transport costs, accommodation costs, where necessary, and other supplies.

#### v) Advocacy, communication and social mobilization

This included activities such as national stakeholder forums, national and county advocacy and launches, media advertisements, information, education and communication material. Further, community mobilization activities with opinion leaders, through roadshows, and with public address systems were also included. The costs for the aforementioned activities included conference packages, per diem allowances for all staff involved, fuel, advertisement production, advertisement truck and generator hire, printing, salaries, and other supplies

#### vi) Data management, monitoring and supervision

At a national level this included activities to support national coordinators and supervisors, policy level oversight, microplanning, preparedness assessments, financial oversight, monitoring and evaluation strengthening, national level support, coordination and planning. At a county level this included activities to support SCHMT supervision, county health management team supervision and microplanning. Costs associated with monitoring and supervision activities included per diem and allowances, lunch, fuel, monitoring tools, ICT equipment maintenance, District Health Information Software 2 (DHIS2) customization, evaluation costs, stationaries, salaries, and other supplies. It also included the electronic logistics management information system that is used by government to manage stocks and immunization. Specifically, the system development costs, cloud hosting service charges, tablets used in facilities, and the messaging services to clients.

#### vii) Vaccine administration costs

The service delivery activities included costs associated with vaccine administration in health facilities. These costs included the time and resources needed for each vaccination, including the preparation for vaccine administration, actual vaccination, and post-vaccination reporting. The costs included staff salaries, supplies and building space for vaccine administration. Outreach costs were also included and assumed 25% of the identified facilities would conduct at least two outreaches per week in the second half of the year. These outreach costs included salaries, transport, and lunch costs.

### Data collection

Resource use data of the identified inputs classified under each sub-activity and cost component were collected through document reviews of national and county level expenditure plans for COVID-19 deployment and structured interviews with purposefully selected national and county level officers involved in the COVID-19 vaccine deployment. Specifically, we interviewed 4 officers from the National COVID-19 vaccine deployment taskforce, and 8 officers involved in county deployment and administration of COVID-19 vaccines drawn from purposely selected, geographically diverse, counties (Kilifi county, and Kakamega county). All identified inputs for the different cost components were costed onto an excel-based tool. Costing assumptions and inputs were agreed upon by the study team and validated by study team members supporting the delivery of COVID vaccines in Kenya.

### Data classification and analysis

First, costs were classified as recurrent versus capital costs. Recurrent costs included resources that are expected to be replaced/consumed within one year. These included costs related to vaccine and related supplies, personnel, transport, all supplies, operations and maintainace, meetings, monitoring and supervision activities, advocacy activities, social mobilization activities. Capital costs on the other hand included the value of resources that provide services for more than one year. These included the costs of equipment, vehicles, building space, and initial training, development costs of the electronic logistics management information system, and the related cloud hosting costs. Equipment and vehicles were assumed to be used for other vaccines as well and therefore their annuitized costs were apportioned based on total EPI and COVID-19 vaccines given in one year.

Second, costs were also classified as either financial or economic costs. Financial costs only reflect the actual costs incurred by the government of Kenya for COVID-19 vaccines and its delivery. For instance, financial costs excluded the vaccine costs procured by COVAX facility and those donated through bilateral negotiations, salaries of all staff involved, volunteer costs, and development and cloud hosting of the electronic logistics management information system. On the other hand, economic costs reflect the opportunity costs and covers the value of all resources used including those not captured in financial outlays by estimating their value. Therefore, economic costs included all financial costs as well as the value of donated vaccines, volunteer time, and staff salaries. For each cost component, both financial and economic costs are estimated.

All costs were valued using purchase prices from the financial records, in few instances where this was not available, the replacement costs from similar items was used. Volunteer costs were valued based on the minimum wage in Kenya for a general labourer [14]. All capital costs were annuitized using a discount rate of 3% over their useful life. Costs were presented in 2021 US dollars. The 2021 average exchange rate from January to August (time point of analysis) was used 1USD=KES 107.87[15].

### Sensitivity Analysis

A univariate sensitivity analysis was done to determine the robustness of the unit cost estimates with variations in key parameters on the economic costs. First, the base costs were varied to range a minimum of $3 to a maximum of $10 assuming that the government of Kenya procures vaccines from Africa Union at different prices [16]. Second, given the dynamic nature and novelty of the disease, training was classified as recurrent costs in the sensitivity analysis; assuming intensive training may need to be given yearly. Lastly, we varied the number of targeted populations receiving 2 doses of the vaccine to 50% receiving 2 doses and the other 50% receiving 1 dose within the one-year time period.

### Ethical approval

The study received ethical approval from the Kenya Medical Research Institute Scientific and Ethics Review Unit; reference KEMRI/SERU/CGMRC-C/4244

## RESULTS

### Number of people to be vaccinated under different scenarios

A total of 8.0 million, 13.4 million, 18.7 million, and 26.8 million adults (above the age of 18) from the Kenyan population are targeted for vaccination under the 30% coverage, 50% coverage, 70% coverage, and 100% coverage vaccination strategies respectively. This is reported in Table 2.

**Table 2:** Number of people to be vaccinated and number of doses administered in a one-year period.

| Vaccination strategy | Total number of adults vaccinated | Number of doses administered (two doses per person) |
| --- | --- | --- |
| 30% coverage | 8,034,303 | 16,068,606 |
| 50% coverage | 13,390,505 | 26,781,011 |
| 70% coverage | 18,746,707 | 37,493,415 |
| 100% coverage | 26,781,011 | 53,562,021 |

### Total Costs

The reported total costs reflect 2-vaccine doses per person vaccinated. The total financial procurement costs ranged from $24.55 million to $352.74 million while the economic procurement costs ranged from $140.65 million to $468.84 million for the 30% and 100% coverage levels respectively.

The total financial delivery costs ranged from $34.39 million to $87.98 million for the 30% and 100% coverage levels respectively. The total economic delivery costs ranged from $98.18 million to $192.09 million for the 30%and 100% coverage levels respectively.

The total financial costs of both procurement and delivery was estimated at $58.94 million, $168.50 million, $278.62 million, and $440.72 million for the 30%, 50%. 70% and 100% coverage levels respectively. Further, the total economic costs of both the procurement and delivery was estimated at $238.83 million, $345.78 million, $474.26 million, and 660.94 million for the 30%, 50%, 70%, and 100% coverage levels respectively. These total incremental costs of procurement and delivery of COVID-19 vaccines are reported in Table 3 Vaccine procurement costs are the largest cost-driver, accounting for 39% to 79% of total financial costs and 58% to 70% of total economic costs across the different vaccine strategies.

**Table 3:** Total incremental procurement and delivery costs for COVID-19 vaccines.

|  | Financial Costs |  |  |  |  | Economic costs |  |  |  |
| --- | --- | --- | --- | --- | --- | --- | --- | --- | --- |
|  | 30% Coverage | 50% coverage | 70% coverage | 100% coverage |  | 30% Coverage | 50% coverage | 70% coverage | 100% coverage |
| | Cost (2021 US\$) | Cost (2021 US\$) | Cost (2021 US\$) | Cost (2021 US\$) | | Cost (2021 US\$) | Cost (2021 US\$) | Cost (2021 US\$) | Cost (2021 US\$) |
| Vaccine procurement costs | 23,219,671.75 | 116,098,358.76 | 208,977,045.77 | 348,295,076.28 |  | 139,318,030.51 | 232,196,717.52 | 325,075,404.52 | 464,393,435.03 |
| Vaccine-related supplies procurement costs | 1,333,472.58 | 2,222,454.30 | 3,111,436.01 | 4,444,908.59 |  | 1,333,472.58 | 2,222,454.30 | 3,111,436.01 | 4,444,908.59 |
| **Total Procurement Costs** | **24,553,144.33** | **118,320,813.05** | **212,088,481.78** | **352,739,984.87** |  | **140,651,503.09** | **234,419,171.81** | **328,186,840.54** | **468,838,343.63** |
| Vaccine supply chain costs | 15,994,252.61 | 26,214,841.49 | 36,352,407.95 | 51,482,054.62 |  | 16,062,882.38 | 26,304,270.46 | 36,461,352.30 | 51,639,446.06 |
| Vaccine safety monitoring & AEFI management costs | 918,838.18 | 1,214,767.59 | 1,510,645.87 | 2,093,067.08 |  | 2,766,280.17 | 3,071,950.08 | 3,377,568.86 | 3,974,600.84 |
| Training costs | 1,056,508.47 | 1,056,508.47 | 1,056,508.47 | 1,056,508.47 |  | 3,331,273.32 | 3,331,273.32 | 3,331,273.32 | 3,331,273.32 |
| Advocacy, communication and social mobilization costs | 6,408,552.56 | 9,180,856.15 | 11,953,159.74 | 16,111,615.12 |  | 28,141,828.72 | 39,348,358.66 | 50,554,888.60 | 67,364,683.51 |
| Data management, monitoring & supervision costs | 7,302,567.20 | 8,243,806.51 | 9,251,213.96 | 10,434,591.74 |  | 31,101,343.73 | 11,684,751.26 | 12,740,132.56 | 13,995,471.10 |
| Vaccine administration costs | 2,708,609.33 | 4,272,376.54 | 6,406,394.28 | 6,801,359.16 |  | 16,779,711.80 | 27,622,495.99 | 39,609,559.53 | 51,792,109.51 |
| **Total Delivery Costs** | **34,389,328.35** | **50,183,156.75** | **66,530,330.27** | **87,979,196.19** |  | **98,183,320.13** | **111,363,099.77** | **146,074,775.17** | **192,097,584.35** |
| **TOTAL COSTS (Procurement and Delivery)** | **58,942,472.68** | **168,503,969.81** | **278,618,812.05** | **440,719,181.05** |  | **238,834,823.22** | **345,782,271.58** | **474,261,615.71** | **660,935,927.97** |

When all procurement costs are excluded, and only delivery costs are considered; the major financial cost driver include vaccine supply chain activities accounting for 47% to 59% of total financial delivery costs. This is then followed by advocacy, communication and social mobilization activities that are estimated at 18-19% of total financial delivery costs and data management, monitoring &supervision activities that are 12% to 21% of total financial delivery costs. However, for economic delivery costs, the main cost driver is advocacy, communication and social mobilization activities accounting for 29% to 35% of total economic delivery costs. This is then followed by supply chain activities, and vaccine administration activities that account for 16% to 27% and 17% to 27% of total economic delivery costs respectively. Across both financial and economic costs, training activities, and vaccine safety monitoring & AEFI management activities were the least cost drivers for the delivery costs. This is illustrated in Figure 2.

**Figure 2:**
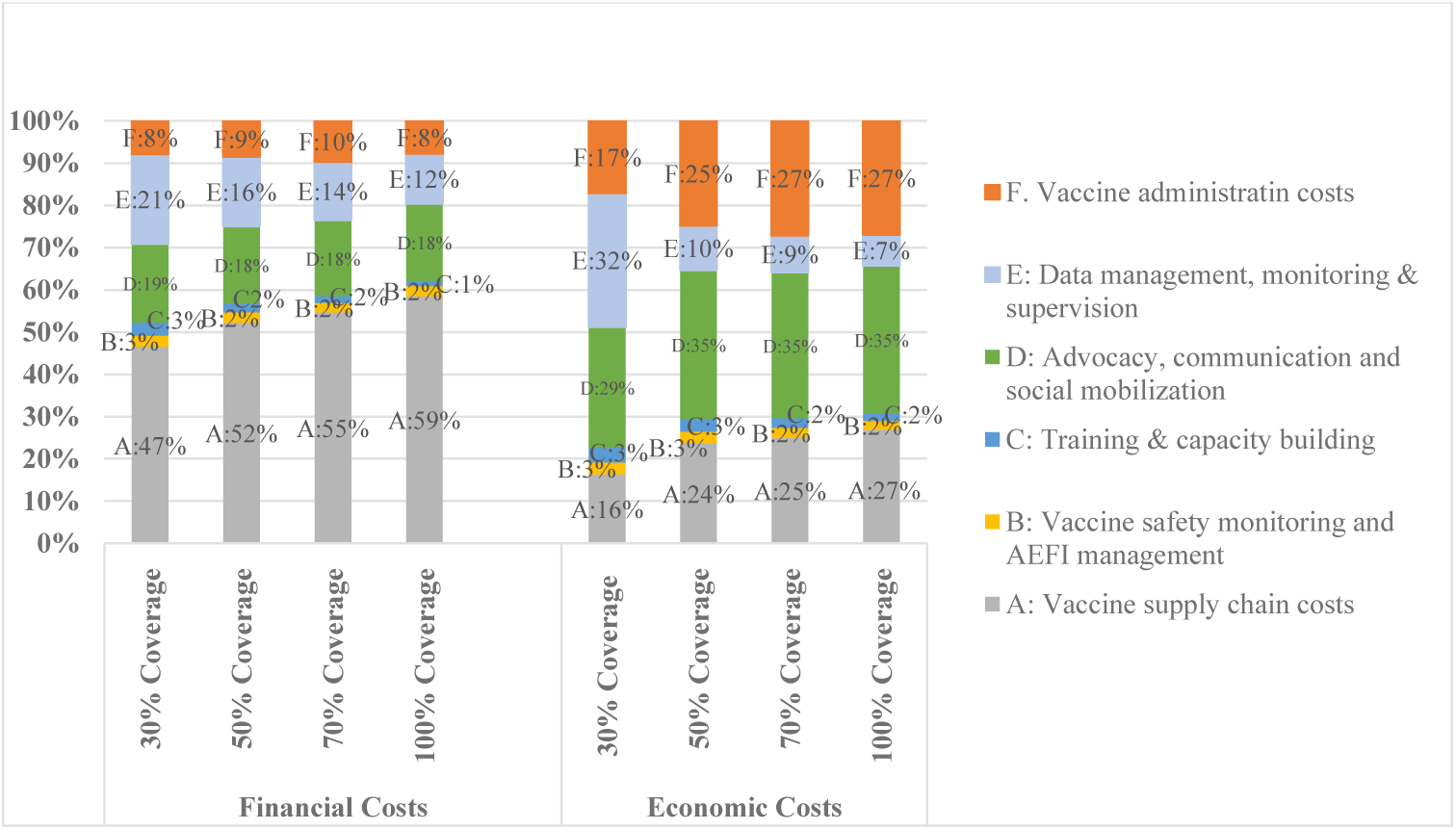
Proportion of cost components contributing to vaccine delivery costs.

### Unit costs

The financial vaccine procurement unit costs to be incurred by the government of Kenya was estimated at $2.89, $8.67, $11.15, and $13.01 per person vaccinated with two-doses for the 30%, 50%, 70% and 100% coverage levels respectively. While the full economic costs of vaccination procurement per person vaccinated with 2 doses was $17.34.

The financial costs of vaccine delivery per person vaccinated with two-doses ranged from $3.29 to $4.28 and the economic costs of vaccine delivery per person vaccinated with two-doses ranged from $7.17 to $12.22 in the 100% and 30% coverage levels respectively.

The total financial unit costs per person vaccinated with two doses (procurement and delivery costs) ranged from $7.34 to $16.47 and the total economic unit costs per person vaccinated with two doses ranged from $29.73 to $24.68 in the 30% and 100% coverage levels. The unit costs are reported in Table 4.

**Table 4:**
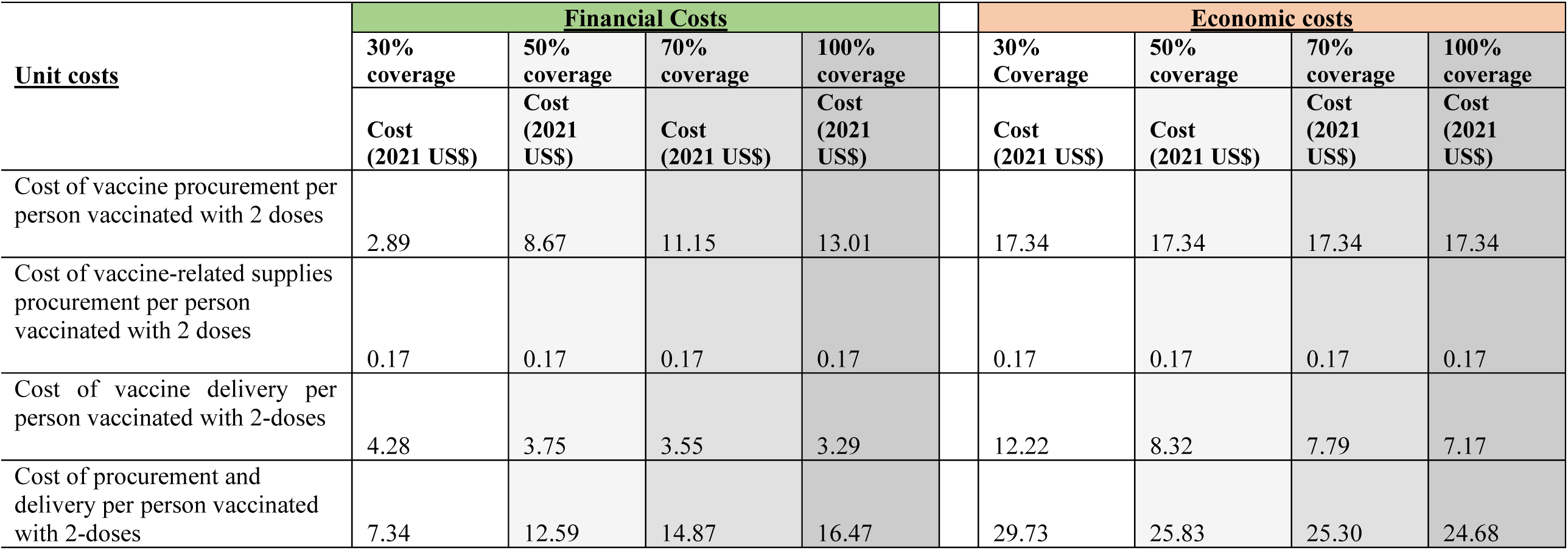
Unit costs.

### Sensitivity analysis

The economic cost of vaccine procurement per person was sensitive to changes in vaccine prices, with a 56% decrease ($7.60) and a 44% increase ($24.94) in the unit cost. The cost of vaccine delivery per person were robust to the changes in the parameters. The economic vaccine delivery per person increased and decreased by less than 10% across all vaccination strategies. This is illustrated in Figure 3.

**Figure 3:**
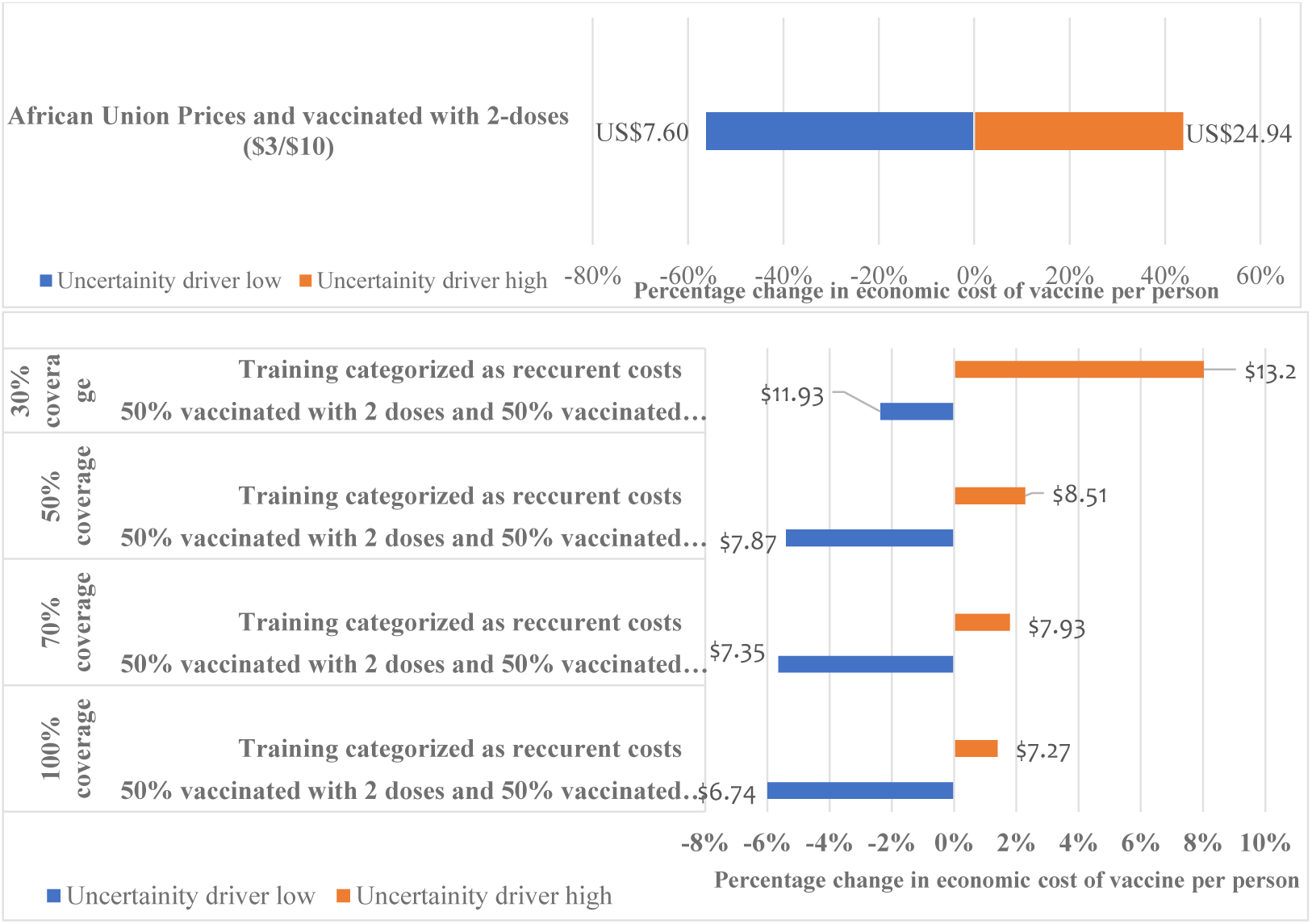
Sensitivity analysis of vaccine costs per person and the cost of vaccine delivery per person across different parameter variations.

## DISCUSSION

We estimated the costs of procuring and delivering the COVID-19 vaccine in Kenya under four different vaccination coverage scenarios. This study provides insights into country-specific unit costs and cost drivers of COVID-19 vaccine delivery costs that can be useful for planning and budgeting purposes and contributes to evidence of costing studies of COVID-19 vaccine delivery.

The largest costs driver was the costs to procure vaccines which contributed 39% to 79% of total financial costs and 58% to 70% of total economic costs across the different vaccination strategies. This is consistent with other vaccine costing studies for other diseases; For instance, a multi-country study done across Ghana, Kenya, and Malawi estimated that the costs of the malaria vaccine were up to 90% of the total financial and economic costs of continuing malaria vaccination after piloting [17]. The sensitivity analysis illustrated that the procurement costs of vaccines per person were sensitive to changes in vaccine prices illustrating the importance of procuring the COVID-19 vaccine at the lowest possible cost. Further, the government may consider policy changes to lower some of the import taxes resulting in a lower financial cost incurred by them.

This study estimates the financial costs of COVID-19 vaccine delivery per person at US$4.28, US$3.75, US$3.55, and US$3.29 for the 30%, 50%, 70% and 100% coverage vaccination strategies. Further, the reported economic costs of COVID-19 vaccine delivery range from US$7.17 to US$12.22 across the vaccination strategies. As expected, the economic costs are consistently higher than the financial costs due to the inclusion of opportunity costs such as salaries, volunteer labour, donated goods, and building space and the development and cloud hosting of the electronic logistics management information system. The estimated unit costs in this study also indicate possible economies of scale in COVID-19 vaccine delivery as the target coverage increases.

Although not directly comparable, a costing study in 92 countries estimated the financial COVID-19 delivery costs of US$3.70 per person vaccinated with two doses for a 20% target population [13]. Our estimated financial cost of COVID-19 delivery per person vaccinated with two-doses is within the estimated range of the other study, however some differences in the costing approaches exist. For instance, Griffiths, et.al [13] assumed the strategy for reaching the elderly and healthcare workers was through outreaches and fixed site respectively and included global and regional level costs. Although our study also assumed both fixed site and outreaches, it only included country level cost estimates, adapted from Kenya’s-specific planned deployment activities and are therefore country-specific. Further, due to existing price differences between countries and different assumed coverage levels, these studies are not directly comparable.

Our findings demonstrated that when considering the incremental financial costs for COVID-19 vaccine delivery, supply chain activities followed by advocacy, communication and social mobilization, and data management, monitoring and supervision were the top three cost drivers. However, for economic costs, vaccine administration costs during service delivery was the third cost driver in addition to supply chain and advocacy, communication and social mobilization. Previous evidence, that focused on total costs reported supply chain costs to be lower than immunization services for EPI vaccines in Kenya [18]. For malaria vaccine delivery in Kenya, evidence suggests that the recurrent cost drivers are social mobilization, service delivery, and monitoring and evaluation [17]. Training although expected to be a cost driver in our study, was among the least cost drivers in both the financial and economic vaccine delivery costs. This was as a result of classifying training as a start-up costs in line with the Global Health Cost Consortium guidelines[19] and assuming that the initial training would last for 2 years.

This study’s limitation is that the study relied on expenditure plans rather than actual expenditures. However, this was mitigated by augmenting this data with structured interviews with vaccine deployment implementors. The strength of this paper is that the assumptions, unit prices, and vaccination scenarios used in the cost analysis are country-specific and therefore can be used to inform local policy and may further inform parameters used in cost-effectiveness models. However, although the results are less generalizable to other similar low-and middle-income settings in the current format, they could potentially be adapted and adjusted to country-specific assumptions.

## Data Availability

All data produced in the present work are contained in the manuscript

## Contributions

EB, AN and JO conceptualized the study. EB, SO and AK collected the data. SO, and EB analysed the data. SO wrote the first draft of the manuscript. All authors contributed to subsequent revisions of the manuscript.

## Patient Consent for publication

Consent to publish findings of the study was obtained from the participants of the study

## Data availability statement

All data relevant to the study are included in the article

## Acknowledgements

We would like to acknowledge the Ministry of Health of the Government of Kenya, and the national COVID-19 vaccine deployment task force for their support in availing the data used for this cost analysis.

